# A mixture model to estimate SARS-CoV-2 seroprevalence in Chennai, India

**DOI:** 10.1101/2022.02.24.22271002

**Authors:** Matt D.T. Hitchings, Eshan U. Patel, Rifa Khan, Aylur K. Srikrishnan, Mark Anderson, K.S. Kumar, Amy P. Wesolowski, Syed H. Iqbal, Mary A. Rodgers, Shruti H. Mehta, Gavin Cloherty, Derek A.T. Cummings, Sunil S. Solomon

**Affiliations:** Department of Biostatistics, College of Public Health & Health Professions, University of Florida; Gainesville, FL, USA; Department of Epidemiology, The Johns Hopkins Bloomberg School of Public Health; Baltimore, MD, USA; YR Gaitonde Centre for AIDS Research and Education (YRGCARE); Chennai, Tamil Nadu, India; Abbott Laboratories; Abbott Park, IL, USA; Department of Biology, University of Florida; Gainesville, FL, USA; Emerging Pathogens Institute, University of Florida; Gainesville, FL, USA

**Keywords:** COVID-19, serosurvey, India, seroprevalence, mixture model

## Abstract

**Background:** Serological assays used to estimate SARS-CoV-2 seroprevalence rely on manufacturer cut-offs established based on more severe early cases who tended to be older.

**Methods:** We conducted a household-based serosurvey of 4,677 individuals from 2,619 households in Chennai, India from January to May, 2021. Samples were tested for SARS-CoV-2 IgG antibodies to the spike (S) and nucelocapsid (N) proteins. We calculated seroprevalence using manufacturer cut-offs and using a mixture model in which individuals were assigned a probability of being seropositive based on their measured IgG, accounting for heterogeneous antibody response across individuals.

**Results:** The SARS-CoV-2 seroprevalence to anti-S and anti-N IgG was 62.0% (95% confidence interval [CI], 60.6 to 63.4) and 13.5% (95% CI, 12.6 to 14.5), respectively applying the manufacturer’s cut-offs, with low inter-assay agreement (Cohen’s kappa 0.15). With the mixture model, estimated anti-S IgG and anti-N IgG seroprevalence was 64.9% (95% Credible Interval [CrI], 63.8 to 66.0) and 51.5% (95% CrI, 50.2 to 52.9) respectively, with high inter-assay agreement (Cohen’s kappa 0.66). Age and socioeconomic factors showed inconsistent relationships with anti-S IgG and anti-N IgG seropositivity using manufacturer’s cut-offs, but the mixture model reconciled these differences. In the mixture model, age was not associated with seropositivity, and improved household ventilation was associated with lower seropositivity odds.

**Conclusions:** With global vaccine scale-up, the utility of the more stable anti-S IgG assay may be limited due to the inclusion of the S protein in several vaccines. SARS-CoV-2 seroprevalence estimates using alternative targets must consider heterogeneity in seroresponse to ensure seroprevalence is not underestimated and correlates not misinterpreted.

## Introduction

Population-based SARS-CoV-2 seroprevalence studies are critical tools for informing public health policy around COVID-19 management^1,2^, and to aid our understanding of key epidemiological parameters of SARS-CoV-2 variants^3^, global burden of the pathogen^4^, and dynamics of immunity^5^. The primary tool for measuring population-level humoral immunity to SARS-CoV-2 has been serological assays that quantify antibody response to components of the virus, indicating prior infection or vaccination. However, most available assays used sera from hospitalized patients proximal to RT-PCR-confirmed SARS-CoV-2 infection to establish thresholds for seropositivity^6,7^. Antibody responses have been found to vary by age, severity of illness, and sex^8,9^. Consistent with other human coronaviruses^10^, waning of antibody levels has been observed for SARS-CoV-2^11,12^ that varies by assay^13,14^, leading to a failure to detect prior infection even in individuals who initially mounted a robust antibody response. As a result, serosurveys using available assays may underestimate cumulative incidence in the general population.

The widespread deployment of vaccines for COVID-19 has further complicated the use of serosurveys to quantify prior SARS-CoV-2 infection. Many authorized COVID-19 vaccines target the spike (S) protein of SARS-CoV-2 promoting the production of anti-Spike immunoglobulin-G (anti-S IgG); consequently, the role of anti-S IgG assays to determine prior infection is now limited. Quantifying antibody response targeting other regions such as the nucleocapsid (N) protein is therefore critical to understanding the burden of infection, and factors associated with infection and re-infection. However, some of these assays appear to be more prone to waning over time compared to those measuring anti-S IgG, thereby underrepresenting true cumulative population-level exposure^15^.

We conducted a large, population-representative serosurvey for SARS-CoV-2 antibody in the south Indian city of Chennai, India (Fig. 1A), using two serological assays to detect antibodies targeting the spike and nucelocapsid proteins of SARS-CoV-2. Samples were collected between January and May 2021, coinciding with the beginning of the Delta variant surge in India (March – July 2021) (Fig. 1B). To investigate the effect of multiple sources of variation in antibody responses on estimates of seroprevalence and its association with individual level factors, we estimated seroprevalence using manufacturer’s cut-offs and compared results to those from a Bayesian mixture model accounting for heterogeneous antibody response between individuals.

**Fig. 1.**
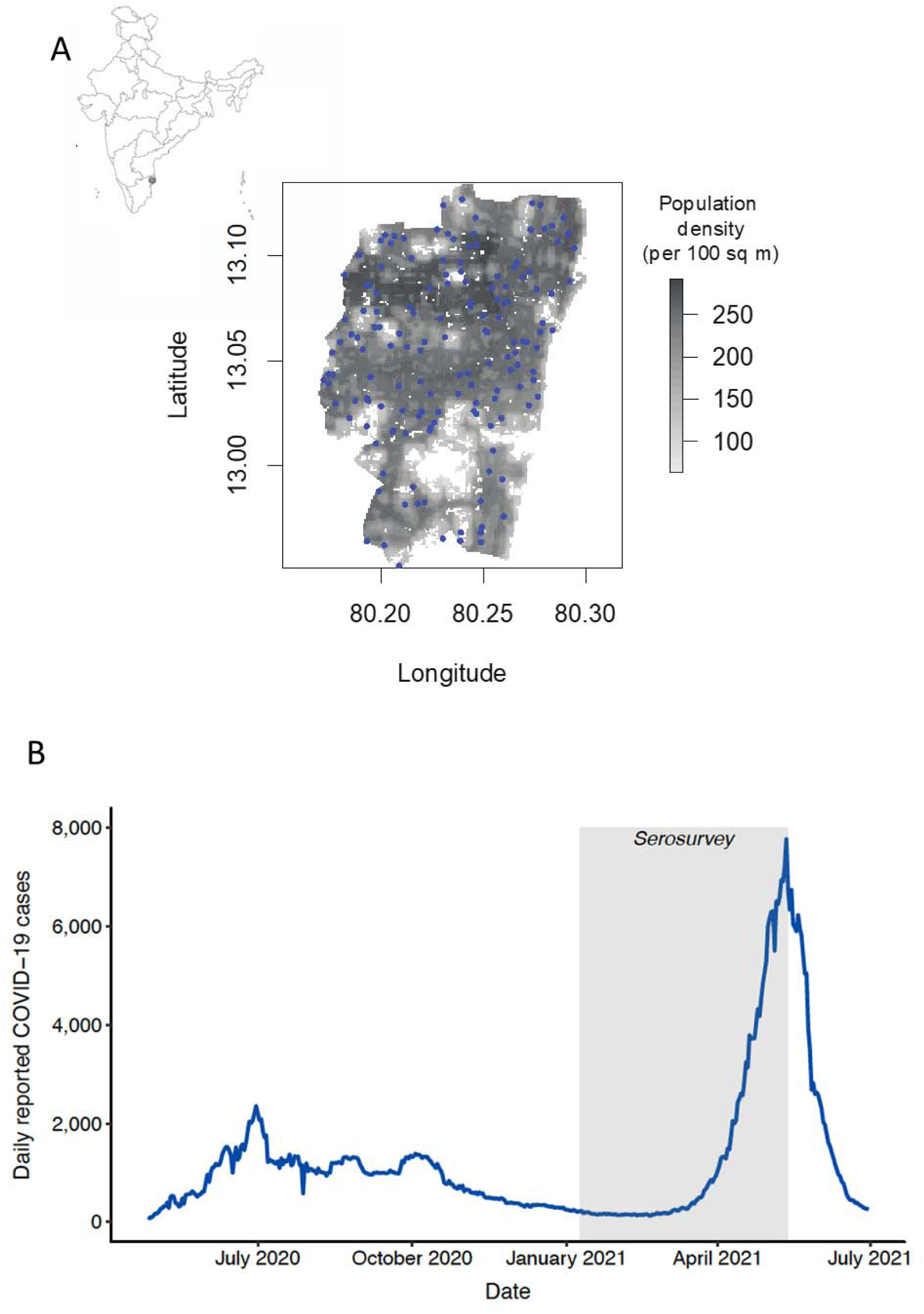
Location of study site (panel A, inset map of India, gray dot) and location of sampling sites in the Chennai Corporation (panel A, blue dots), and daily reported COVID-19 case counts in Chennai, India (panel B) with serosurvey sampling window (shaded gray area).

## Results

### Study population

A total of 2,723 households were recruited across 103 spatial locations randomly selected using a probability proportional to population size approach to cover 12 pre-defined zones within the greater Chennai corporation (Fig. 1A). The median household size was 3 (IQR, 2-4). A median of 1 person (interquartile range [1-2]) was recruited per household for a total sample size of 4,828 eligible persons. Of 4,813 participants recruited who provided a blood specimen, 4,809 were tested for anti-N IgG and/or anti-S IgG among whom 4,677 from 2,619 households had complete data on covariates of interest and comprise the analytical sample (Fig. S1).

In the analytical sample, the median age was 38 years (IQR, 26-51 years). 298 (6.4%) were less than 18 years of age and 28% were 50 years of age or older; about half (48.6%) the study population were female (Table 1). Nearly 20% lived in a household where no household member had completed high school and 67% experienced income loss due to the COVID-19 pandemic. Sixty nine percent lived in households with fans only. At the time of the survey, 11.4% reported receiving at least one dose of a COVID-19 vaccine.

**Table 1.** Characteristics of the study population.

| Study characteristic | n (%)<br>(N=4,677) |
| --- | --- |
| Study period |  |
| 9 Jan - 14 Feb | 1,259 (26.9) |
| 15 Feb - 9 Mar | 792 (16.9) |
| 10 Mar - 24 Mar | 820 (17.5) |
| 25 Mar - 19 Apr | 894 (19.1) |
| 20 Apr - 13 May | 912 (19.5) |
| Age group (in years) |  |
| 5-17 | 298 (6.4) |
| 18-29 | 1,208 (25.8) |
| 30-39 | 937 (20.0) |
| 40-49 | 917 (19.6) |
| ≥50 | 1,317 (28.2) |
| Gender |  |
| Female | 2,272 (48.6) |
| Male | 2,394 (51.2) |
| Transgender | 11 (0.2) |
| Maximum household education |  |
| No formal education | 84 (1.8) |
| Standard 1-8 | 859 (18.4) |
| 9th standard or more | 3,734 (79.8) |
| Household income |  |
| No change/increase due to COVID-19 | 1,551 (33.2) |
| Negative COVID-19 impact | 3,126 (66.8) |
| Household ventilation system |  |
| Neither coolers nor air conditioning | 3,232 (69.1) |
| Coolers only | 268 (5.7) |
| Air conditioning only | 986 (21.1) |
| Air conditioning and cooling systems | 191 (4.1) |
| Median household crowding index (person/rooms), (IQR) | 2.0 (1.5-3.0) |
| Median no. of regular visitors to home, (IQR) | 0 (0-2) |
| Received COVID-19 vaccine |  |
| No | 4,142 (88.6) |
| Yes | 535 (11.4) |

\*Data are sample sizes and corresponding column percentages (%), unless noted otherwise

### SARS-CoV-2 anti-N and anti-S IgG seroprevalence

The seroprevalence of anti-S IgG using the manufacturer’s cut-off of 50 AU/ml was 62.0% (95% confidence interval [CI], 60.6 to 63.4). For the anti-N IgG, the seroprevalence using the manufacturer’s normalized index cut-off of 1.4 was substantially lower at 13.5% (95% CI, 12.6 to 14.5). Among participants who reported never having received a COVID-19 vaccine in the past, the prevalence of anti-S and anti-N IgG was 60.5% (95% CI, 59.0 to 62.0) and 12.9% (95% CI, 11.9 to 13.9), respectively.

In the mixture model, the population was assumed to be divided into individuals who were seronegative or seropositive. The distribution of measured IgG across the sample was used to infer the proportion of the population that came from the seropositive compartment. The mixture model is illustrated in Fig 3A and 3B, demonstrating the two compartments and the overall fit to the data. A log-normal distribution was chosen to model the seroresponse among seropositive individuals for both assays after a visual inspection of the model fit to the data by age and time quintile (Figs. S2 and S3). A log-logistic distribution was chosen to model the assay variability among 580 samples collected in 2016-2017 (pre-pandemic), to represent the distribution of IgG among seronegative individuals (Fig. S4). The model-estimated seropositivity probability is plotted against anti-N and anti-S IgG in Fig. S5. Using the mixture model-based determination of seropositivity, the estimated seroprevalence of anti-S and anti-N IgG was 64.9% (95% Credible Interval [CrI], 63.8 to 66.0) and 51.5% (95% CrI, 50.2 to 52.9), respectively. When the analysis was restricted to individuals who self-reported that they had not received a COVID-19 vaccine, seroprevalence using the mixture model approach was 63.7% for anti-S IgG (95% CrI, 62.6 to 64.8) and 53.4% for anti-N IgG (95% CrI, 51.9 to 54.8).

Agreement between the two assays using the manufacturer’s cut-offs was low (Cohen’s Kappa 0.15; 95% CI 0.14, 0.17). Defining an individual as seropositive if the mixture model-estimated seropositivity probability was ≥0.5, the agreement was substantially better (Cohen’s Kappa 0.66; 95% CI 0.63, 0.68). To understand factors underlying the difference in seroprevalence between the assays, we estimated the anti-S to anti-N relative seroprevalence over time and age among individuals reporting no COVID-19 vaccine receipt (Table S1). Anti-S seroprevalence increased relative to anti-N seroprevalence from late March 2021 onward, but there was no clear trend with respect to antibody response and age.

### Factors associated with SARS-CoV-2 seroprevalence by assay and statistical method

Using the manufacturer’s cut-offs, multivariable logistic regression suggested contrasting associations between age and seropositivity for each assay, with increased odds of anti-N IgG seropositivity among older individuals (adjusted odds ratio (aOR) for ≥50 years vs. 18-29 years, 2.35; 95% CI, 1.76 to 3.13) but no relationship between age and anti-S IgG seropositivity (aOR for ≥50 years vs. 18-29 years, 0.95; 95% CI, 0.77 to 1.18; Table 2). Moreover, the relationship between some indicators of socioeconomic status and seropositivity differed between the two assays. For example, better ventilation was associated with higher anti-N seropositivity (aOR for air conditioning and fans in the home vs. fans only 1.43; 95% CI, 1.08 to 1.89) but lower anti-S seropositivity (aOR for air conditioning, cooling systems, and fans vs. fans only 0.48; 95% CI, 0.30 to 0.76). There was no association between the number of regular outside visitors to the home and anti-N seropositivity (aOR per household visitor 0.95; 95% CI, 0.87 to 1.04) while having more outside visitors to the home was significantly associated with anti-S seropositivity (aOR 1.11; 95% CI, 1.03 to 1.19).

**Table 2.**
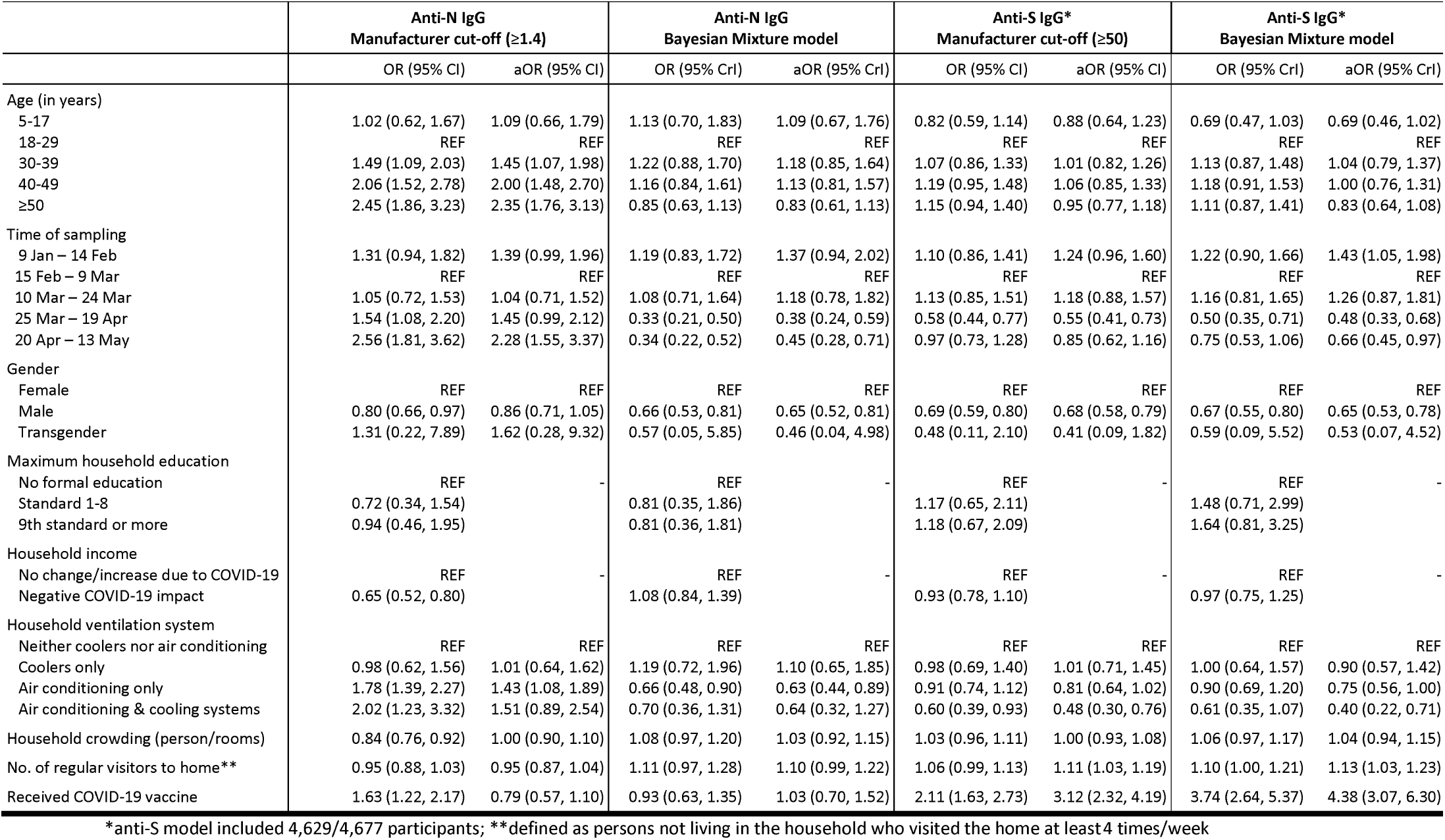
Correlates of anti-N and anti-S seropositivity among 4,677 individuals sampled across Chennai, India: results from multivariable logistic regression using manufacturer’s cut-offs and from a Bayesian mixture model

In the mixture model there was no clear trend in seropositivity by age, although a suggestion of lower anti-N and anti-S seropositivity among older adults (aOR for ≥50 years vs. 18-29 years, 0.83; 95% CrI 0.61 to 1.13 and 0.83; 95% CrI, 0.64 to 1.08, respectively), and lower anti-S seropositivity among children (aOR for 5-17 years vs. 18-29 years, 0.69; 95% CrI, 0.46 to 1.02). Markers of lower socioeconomic status were consistently associated with higher seropositivity in univariable analyses, and household ventilation was associated with lower anti-N and anti-S seropositivity in multivariable analyses (aOR for air conditioning, cooling systems, and fans in the home vs. fans only 0.64; 95% CrI, 0.32 to 1.27 and 0.40; 95% CrI, 0.22 to 0.71 respectively). An increase in the number of regular visitors to the home was associated with higher anti-N and anti-S seropositivity (aOR per additional visitor per week 1.10; 95% CrI, 0.99 to 1.22 and 1.13; 95% CrI, 1.03 to 1.23 respectively). In both assays, men had lower seropositivity compared to women (aOR for men vs. women 0.65; 95% CrI, 0.52 to 0.81 for anti-N and 0.65; 95% CrI, 0.53 to 0.78). Self-report of having received a COVID-19 vaccine was associated with 4.38-fold (95% CrI, 3.07 to 6.30) change in odds of anti-S seropositivity, but was not associated with anti-N seropositivity (aOR 1.03; 95% CrI 0.70 to 1.52).

In the mixture model, the age pattern of mean seroresponse among seropositive individuals was similar for the two assays with all groups having higher seroresponse than individuals 18-29 years of age (Fig. 2 and Table S2); however, credible intervals for children aged 5-17 and adults 30-39 years excluded 1 for the anti-S assay only. The mean seroresponse among seropositive individuals increased over time for both assays, from late March onwards compared to February (Fig. 2 and Table S2), reflecting robust IgG response following infections acquired at the start of the Delta wave.

**Fig. 2.**
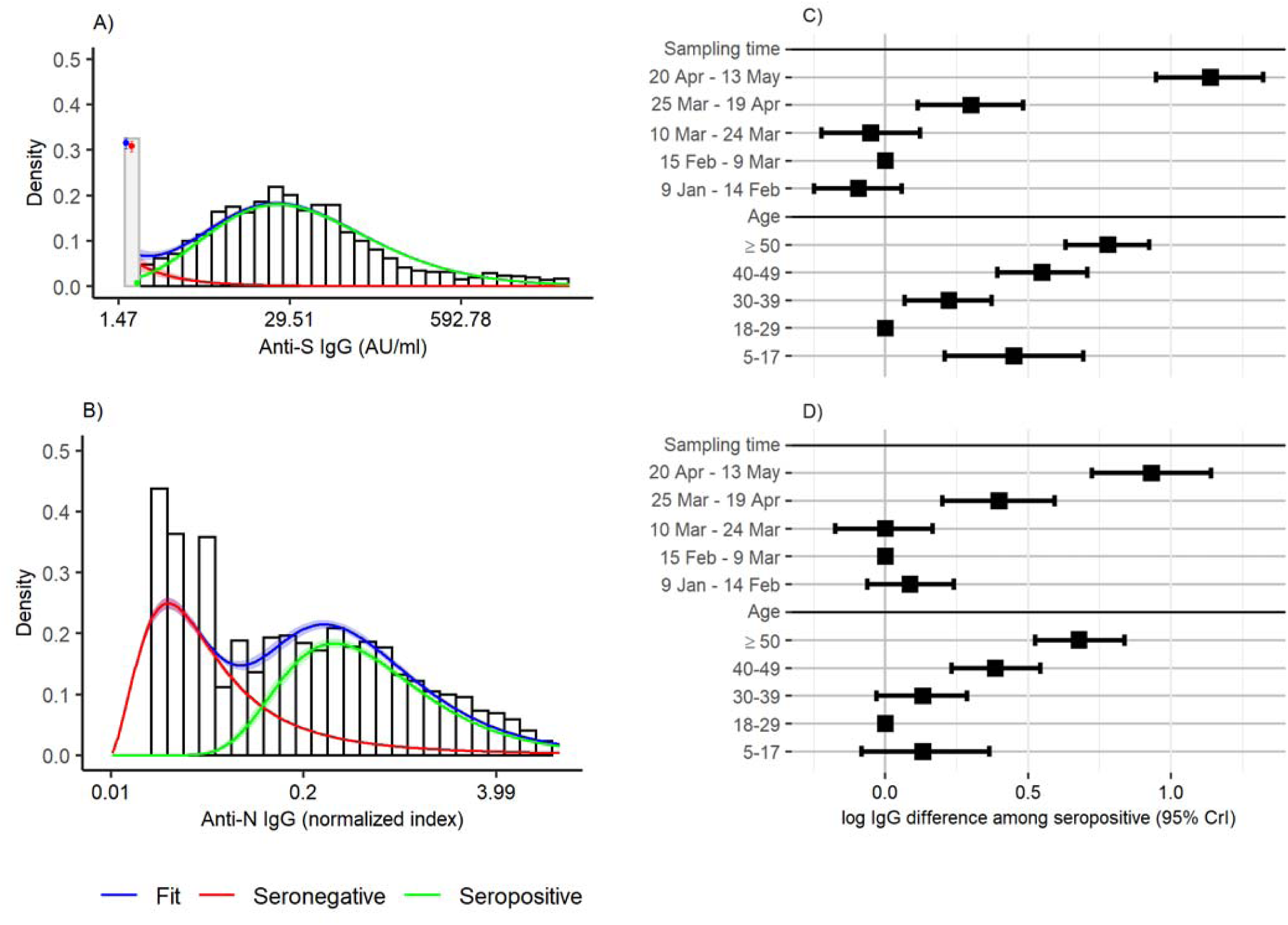
Illustration of mixture model distribution and fit to the anti-S IgG (panel A) and anti-N IgG (panel B) data, and association between age, sampling time, and log IgG among seropositive individuals for anti-S IgG (panel C) and anti-N IgG (panel D). In panels A and B, the bars represent the distribution of IgG in the data. The red, green, and blue lines represent the distribution of IgG values in the seronegative and seropositive compartments and overall, respectively, with shaded bands reflecting 95% CrI for each distribution. In Panel A, the gray bar represents the proportion of samples that were below the LLOQ, while the points represent the probability for each compartment to be below the LLOQ.

## Discussion

In this population-based sample of residents from Chennai, India we observed a strikingly disparate prevalence of SARS-CoV-2 antibodies using two IgG assays (62.0% anti-S IgG seroprevalence vs. 13.5% anti-N IgG seroprevalence) when using manufacturer’s cut-offs. However, using a mixture model to infer seropositivity from the raw antibody response, prevalence of SARS-CoV-2 antibodies from each assay was more similar (64.9% anti-S IgG seroprevalence and 51.5% anti-N IgG seroprevalence). Further, we observed qualitative differences in the associations between key exposures of interest and SARS-CoV-2 anti-N IgG seropositivity when using manufacturer’s cut-offs and the mixture model to determine seropositivity. When applying the manufacturer’s cut-off, we observed higher anti-N IgG seropositivity among older individuals (likely due to their stronger seroresponse) and among individuals with better household ventilation, but not for anti-S IgG. Associations were consistent between assays when applying the mixture model.

The estimated anti-S IgG seroprevalence of 60-65% is higher than the 42.9% seroprevalence measured in Chennai around the same time^16^, but consistent with the 53.8% seroprevalence measured in urban areas of neighboring Karnataka State in August 2020^17^. However, the utility of anti-S based serosurveys to quantify population exposure to SARS-CoV-2 is rapidly dwindling given increasing vaccine coverage in many countries. Currently, most vaccines in use globally target the spike region of SARS-CoV-2 and elicit the production of antibodies to the Spike region. The anti-S IgG surveys cannot distinguish anti-S antibodies produced as a result of vaccination, prior vaccination, or both. Nevertheless, population-based serosurveys remain critical to understanding the burden of infection in a population over time and the role of humoral antibodies from prior infection against re-infection and emerging variants.

The anti-nucleocapsid (anti-N) IgG assay provides one such tool to quantify population-level exposure in the presence of widespread deployment of vaccines targeting the Spike region. However, population serosurveys conducted in other contexts have found discrepancy between anti-N and anti-S IgG seroprevalence (e.g. 38% vs. 64% seroprevalence to anti-N and anti-S IgG in India after the Delta wave)^18^, that persisted among unvaccinated individuals who had lab-confirmed SARS-CoV-2 infection (63% vs. 82%)^18^. In this study we estimated anti-N IgG seroprevalence of 13.5% using the Abbott ARCHITECT assay and associated cut-off, a five-fold difference compared to the anti-S IgG seroprevalence.

Given these limitations, mixture models have been proposed^19^ and used previously to perform inference on serological data^20–22^. A major strength of these approaches is that they infer seropositivity using the distribution of raw antibody values in the population, obviating the need for a strict, binary cut-off. In doing so, these models reduce measurement error from equivocal assay results, and explicitly account for the uncertainty arising from heterogeneity in antibody response between individuals. In this case, use of the mixture model markedly improved the agreement between the assays.

In addition to identifying the burden of infection, serosurveys are also used to identify factors associated with infection to plan appropriate interventions to curb transmission. Due to the association between age, severity, and antibody response, as well as the association between demographic factors, the timing of infection and subsequent waning, a risk factor analysis based on manufacturer’s cut-offs produced erroneous results based on anti-N IgG seropositivity, including a positive association between improved ventilation and seropositivity and higher seropositivity among individuals aged ≥50 years. On the other hand, the mixture model identified the same risk factors for the two assays by explicitly accounting for varying antibody response by age and time of sample collection. Due to the waning of measured antibody levels, factors associated with seroprevalence could be associated with earlier infection times. Identification of such risk factors in different contexts across the globe is crucial for better targeting of public health interventions, and statistical methods to perform these analyses account for characteristics of the assay.

Some discrepancy between anti-N and anti-S IgG seroprevalence persisted even when we applied the mixture model, with 51.5% anti-N IgG seropositivity compared to 64.9% anti-S IgG seropositivity. Possible explanations for this discrepancy include differential waning of anti-N vs. anti-S in the initial months following infection, with anti-N waning faster, and the possibility that some persons may never mount an anti-N response following initial infection. Previous studies have demonstrated faster waning of Abbott anti-N IgG over time since infection compared to other assays^15,23^. In our sample, the difference was observed throughout the time of sample collection and increased slightly towards the end of the sampling period. As incidence increased during sample collection, and antibody response takes weeks to develop, these findings are consistent with increased durability of the anti-S IgG response and/or with a more robust anti-S IgG following an acute infection^24^. Finally, some participants may have also misreported their COVID-19 vaccination status which might contribute to the discrepancies between these assays. Together, these findings suggest that serosurveys using anti-N IgG assays with standard statistical approaches and utilizing manufacturer cut-offs could significantly underestimate true seroprevalence in a community. Further theoretical work should focus on the utility of serosurveys using anti-S and anti-N IgG assays to identify recent and non-recent infections, leveraging the waning of the latter assay.

This study has several limitations. Given the difficulty of implementing a survey amidst an ongoing pandemic, COVID-19 safety protocols prevented higher enrollment per household. This might have impacted the representativeness of the sample. Although our seroprevalence estimates were higher than estimates from others studies conducted in Chennai during this time^16^, they were comparable to similar studies in urban settings across India^17^. In addition, these assays and corresponding antibody waning only relate to the humoral response; more studies are required to examine the association between the humoral response of anti-N and cell-mediated responses. These studies will be critical to the interpretation of anti-N IgG seroprevalence data and to identify thresholds related to protection from future infection. Finally, although we observed decline in seropositivity, we did not correct our seroprevalence estimates to account for such waning; therefore, the seroprevalence in this sample likely underestimates the cumulative incidence.

In summary, a mixture model produced improved estimates of SARS-CoV-2 anti-N IgG seropositivity and identification of factors associated with seropositivity. While estimates of seropositivity based on the anti-S assay were less affected by antibody waning, future seroprevalence studies will be challenged by increasing vaccination and the inability to distinguish prior infection from vaccination-induced immunity. Thus, future work should focus on making methods such as mixture models accessible and computationally feasible for a wide array of researchers undertaking serosurveys for SARS-CoV-2 and other novel pathogens.

## Methods

### Study design

We conducted a cross-sectional, household-based serosurvey to measure the seroprevalence of SARS-CoV-2 antibodies in Chennai, India. The study was approved by the IRB of YR Gaitonde Centre for AIDS Research and Education (YRGCARE) and by the IRB of Johns Hopkins University for secondary data analysis. All participants provided written informed consent.

### Study setting

The south Indian city of Chennai, the capital of the state of Tamil Nadu, is home to an estimated 7.1 million residents with a population density of 26,553 per square kilometer. The city was broadly divided into 12 administrative zones prior to its expansion to 15 zones that increased the city area from 174 sq km to 426 sq km; we included the original 12 zones in our sample. The first case of COVID-19 was reported in Chennai on March 7, 2020 and as of October 20, 2021 had reported a cumulative of 532,529 confirmed cases and 8,187 confirmed deaths^25^.

During the first wave (April through July 2020), test positivity peaked at about 23% with an average of 8,000 tests per day. A complete lockdown was instituted between March 23, 2020 and June 8, 2020 during which period, inter-district movement was restricted and international travel was suspended. The highest single-day case count of 2,509 was observed on June 30, 2020, with a steady decline thereafter. A second surge in COVID-19 infections was observed in Chennai between April through June 2021 with a peak daily case count of 7,772 reported on May 12, 2021 followed by a steady decline^25^. A second lockdown was instituted on May 6, 2021 and was in effect through late June 2021. Prior community-based serosurveys in Chennai estimated the anti-N IgG seroprevalence of SARS-CoV-2 to be 18.4% in July, 2020^26,27^ and 30.1% in October, 2020^27^, and the anti-S seroprevalence to be 40.9% in October, 2020^28^. COVID-19 vaccination commenced in Chennai on January 16, 2021 for health care workers with expansion to persons 60 years of age or older two months later. The only vaccines available at the time in India were Covishield (Serum Institute of India; https://www.seruminstitute.com/product_covishield.php) and Covaxin (Bharat Biotech; https://www.bharatbiotech.com/covaxin.html) both of which target the Spike region.

### Study sample

We selected 150 locations from within the boundaries of Chennai proportional to the population size using WorldPop data from 2020 with a goal of sampling from 100 locations. Of the 150 locations, 123 were determined to be viable (e.g., residential). The target sample size at each location was 50 households. Within each location, the starting point was the household nearest to the designated GPS location. If the starting point identified multiple houses, study staff counted the number of houses and selected randomly from the total number of houses. If the starting point was a high rise or apartment building, the floors were enumerated and a floor was randomly selected for sampling. If there was no household in a specified location, study staff would move systematically from the right of the starting location until finding a home. The goal was to enroll as many individuals 5 years of age and older as possible from each household while ensuring that within each location there were at least 10 individuals in each of three groups (5-25 years, 26-45 years and greater than 45 years). The study team returned to households a maximum of 3 times if an eligible adult was not available. The locations of Chennai and of the sampling sites are displayed in Fig. 1.

### Data collection

In each household, an adult (18 years of age and older) who could speak on behalf of the household was identified as the index. He/she was asked to enumerate all members of the household and provide information on basic sociodemographics, history of COVID-19 symptoms, SARS-CoV-2 testing history and COVID-19 illness history. Index participants were additionally asked questions about sociodemographics, their own lifetime COVID-19 symptoms and testing history, comorbidities and general health care access, adoption of non-pharmaceutical interventions (social distancing and masking) and activities in the prior 2 weeks.

Index members were also asked to recruit other household members who were 5 years of age and older. Household members were asked to respond to a similar series of questions with the exception of the household inventor.

All individuals who participated were offered SARS-CoV-2 antibody testing; samples were collected (6-10 ml of whole blood) prior to conducting the survey. All participants provided written informed consent and were compensated for their time. The protocol was approved by the YR Gaitonde Center for AIDS Research and Education Institutional Review Board.

580 samples that were collected in 2016-17 from clients seeking testing for HIV and related conditions that were available at the YRGCARE laboratory’s specimen repository were tested for antibodies to establish pre-pandemic anti-N and anti-S IgG response.

### Laboratory procedures

All samples (including pre-pandemic specimens) were run on an Abbott ARCHITECT™ *i*1000 instrument and tested with CE marked SARS-CoV-2 IgG and AdviseDx SARS-CoV-2 IgG II assays according to the ARCHITECT operations manual and assay package insert instructions. Briefly, the SARS-CoV-2 IgG assay is an automated Chemiluminescent Microparticle Immunoassay (CMIA) used for the qualitative detection of IgG antibodies directed against the SARS-CoV-2 N-protein. Assay results are measured in Relative Light Units (RLU) and reported as an index value of the ratio of specimen to calibrator RLU signal (S/C). Index values ≥1.4 S/C indicate a SARS-CoV-2 IgG seropositive result. The AdviseDx SARS-CoV-2 IgG II assay is a quantitative automated CMIA used for the quantitative detection of IgG antibodies directed against the receptor binding domain (RBD) of the SARS-CoV-2 S-protein with assay results reported in AU/mL. Assay calibration is performed using 6-point calibration referencing an internal reference standard at each concentration level. Assay linearity was shown between 21.0 and 40,000 AU/mL, with results <50.0 AU/mL reported as negative and ≥50.0 reported as positive.

### Outcomes

The response variables were anti-nucelocapsid (anti-N) and anti-spike (anti-S) IgG, measured in index and AU/ml respectively. For the analysis of seropositivity defined by manufacturer’s cut-offs, the outcome was a binary variable. For the mixture model, we log-transformed the raw values and translated each log-transformed value so that all outcome values were positive.

### Statistical Analysis

For the analysis of seropositivity using manufacturer’s cut-offs, we performed univariable and multivariable logistic regression on seropositivity defined as above. We accounted for clustering by household using generalized estimating equations with an exchangeable correlation structure.

For the mixture model, for each outcome we assumed that the population comprised two unobserved groups defined by their IgG level: those who truly had no exposure to SARS-CoV-2 or response to vaccination (“seronegative”), and those who had some exposure and/or response (“seropositive”). Among the seropositive, the translated, log-transformed IgG followed a defined distribution (normal or log-normal), representing the strength of response to a previous exposure. Among the seronegative, the translated, log-transformed IgG followed a defined distribution (normal or log-logistic), representing assay variability among seronegatives. For the anti-S IgG, the seropositive and seronegative distributions were censored below at the LLOQ.

We derive the likelihood of this model as an extension of Hitchings et al^21^. Let Y=log(IgG)+min(log(IgG)), and let the mean response for an individual *i* be

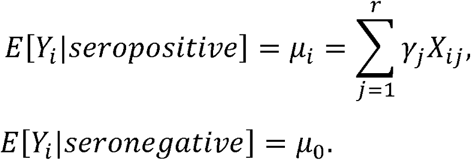

μ_i_ represents the mean of the seropositive distribution with variance/shape parameter σ > 0 and probability density function *f*_*SP*_(*x*), and the γ parameters represent associations between covariates X_ij_ and Y. μ_0_ represents the mean of the seronegative distribution with variance/scale parameter σ_0_ > 0 and probability density function *f*_*SN*_ (*x*). Being seropositive is determined by a Bernoulli distribution with probability p_i_, which links to a different set of covariates X’_ij_ with a logistic model

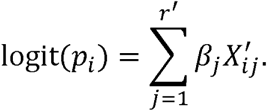

For the anti-N, the log-likelihood of the model given *N* individuals in the serosurvey is

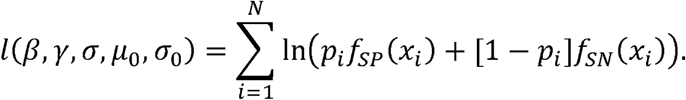

For the anti-S, assuming that the population consists of n individuals with IgG ≥ LLOQ and m individuals with IgG < LLOQ, the log likelihood of the model given the data is

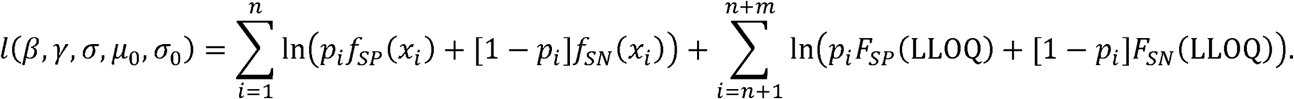

In addition, a household-level random effect was included on the probability of being seropositive. Specifically, a number *h*_*k*_ drawn from a normal distribution with standard deviation α was added to the log-odds of seropositivity for each household, so that the probability of being seropositive *p*_*ik*_ for individual *i* in household *k* is given by

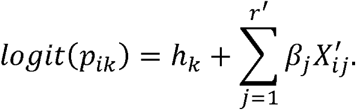

Parameter estimation was done by maximizing the log-likelihood using Bayesian MCMC in the R package rstan^29^, using R, version 3.6.1. Estimates are presented alongside the 95% credible interval estimated from the posterior likelihood.

Finally, data on pre-pandemic negatives were used to inform the distribution of IgG among seronegative individuals. The mean and variance/scale of the negative distribution was estimated using a simplification of the above model in which all individuals were truly seronegative. The median estimated mean and variance/scale parameters were used as the mean of the prior distribution for these two parameters in the full mixture model (using a uniform prior with specified range either side). For other parameters, uniform priors with limits encapsulating range of reasonable values were used for all regression coefficients and the variance/scale of the seroresponse distribution, and a uniform prior on (0,2) was used for α.

Separate models were constructed for anti-S and anti-N positivity. For both the standard logistic regression models and the mixture models, we included age (in quintiles) and time of sample (in quintiles) as explanatory variables. Similarly, both were included to model the mean seroresponse among seropositive individuals. Then, for a chosen set of explanatory variables, we fit univariable models to each outcome separately, including each variable in the log-odds component only. We compared each model to the “null” model (with age and time only) with leave-one-out information criteria (LOOIC) using the loo R package^30^. Each variable that improved the LOOIC of the model was included in a multivariable analysis, with no further model selection performed. For comparability of the two assays, we included any variables that improved the LOOIC of either anti-N or anti-S model, such that the multivariable models contained the same variables. In the mixture model, we used a log-logistic model for the non-response distribution and explored both a normal and log-normal distribution for the seroresponse distribution.

For the mixture model, we estimated overall seroprevalence in the following way: for each of N=10,000 parameter draws from the posterior distribution, we calculated the probability density of seroresponse *f*_*SP*_(*x*)and non-response *f*_*SN*_(*x*) for each individual with IgG *x*. The probability of being seropositive for each individual was *f*_*SP*_(*x*) / (*f*_*SP*_(*x*) + *f*_*SN*_(*x*)).The estimated seroprevalence and 95% CrI were taken as the 2.5^th^, 50^th^, and 97.5^th^ percentiles of the mean seropositive probability across simulations, thus accounting for uncerainty in parameters.

## Data Availability

Data available on reasonable request. Code to perform mixture models available at www.github.com/mhitchings/seromixmodels.

## Author contributions

SSS, SM, and DATC conceived the study. MDTH and EP completed analysis and developed methodology together with SM, DATC, and SSS. RK, AKS, MA, KSK, SHI, MAR, and GC performed the investigation. Project administration was performed by RK and AKS. MDTH, EP, APW, SM, DATC, and SSS wrote the original draft, MDTH, DATC, and EP made visualizations, and all authors reviewed and edited the manuscript. SSS acquired funding. The corresponding author attests that all listed authors meet authorship criteria and that no others meeting the criteria have been omitted.

## Ethics approval

The study was approved by the IRB of YR Gaitonde Centre for AIDS Research and Education (YRGCARE) and by the IRB of Johns Hopkins University for secondary data analysis. All participants provided written informed consent.

## Acknowledgments

This serosurvey was implemented with guidance, support and cooperation from Greater Chennai Corporation, Government of Tamil Nadu. We thank the field teams and all the study participants.

## Supplementary Materials

**Fig. S1.**
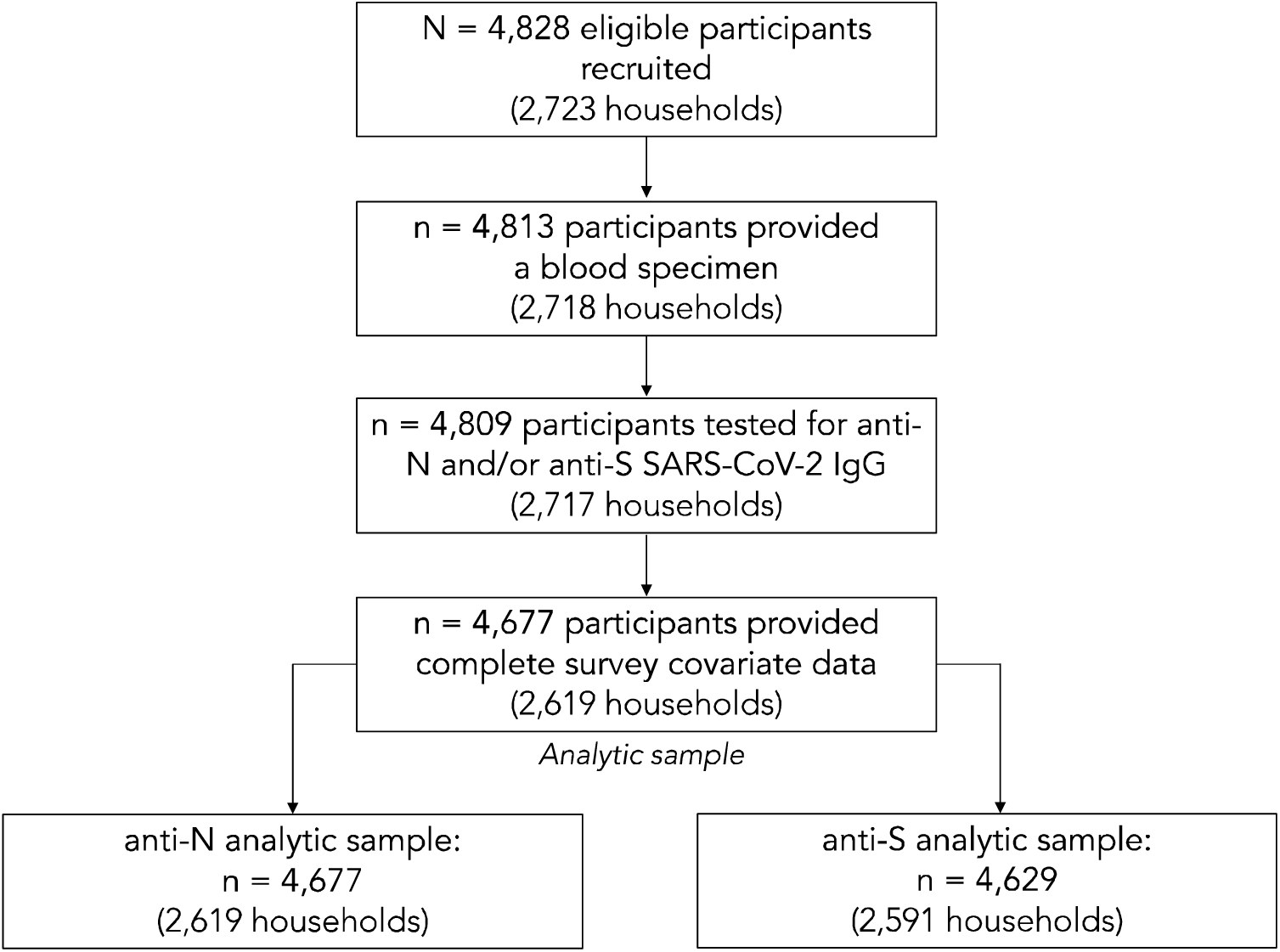
Flowchart of study sample selection

**Fig. S2.**
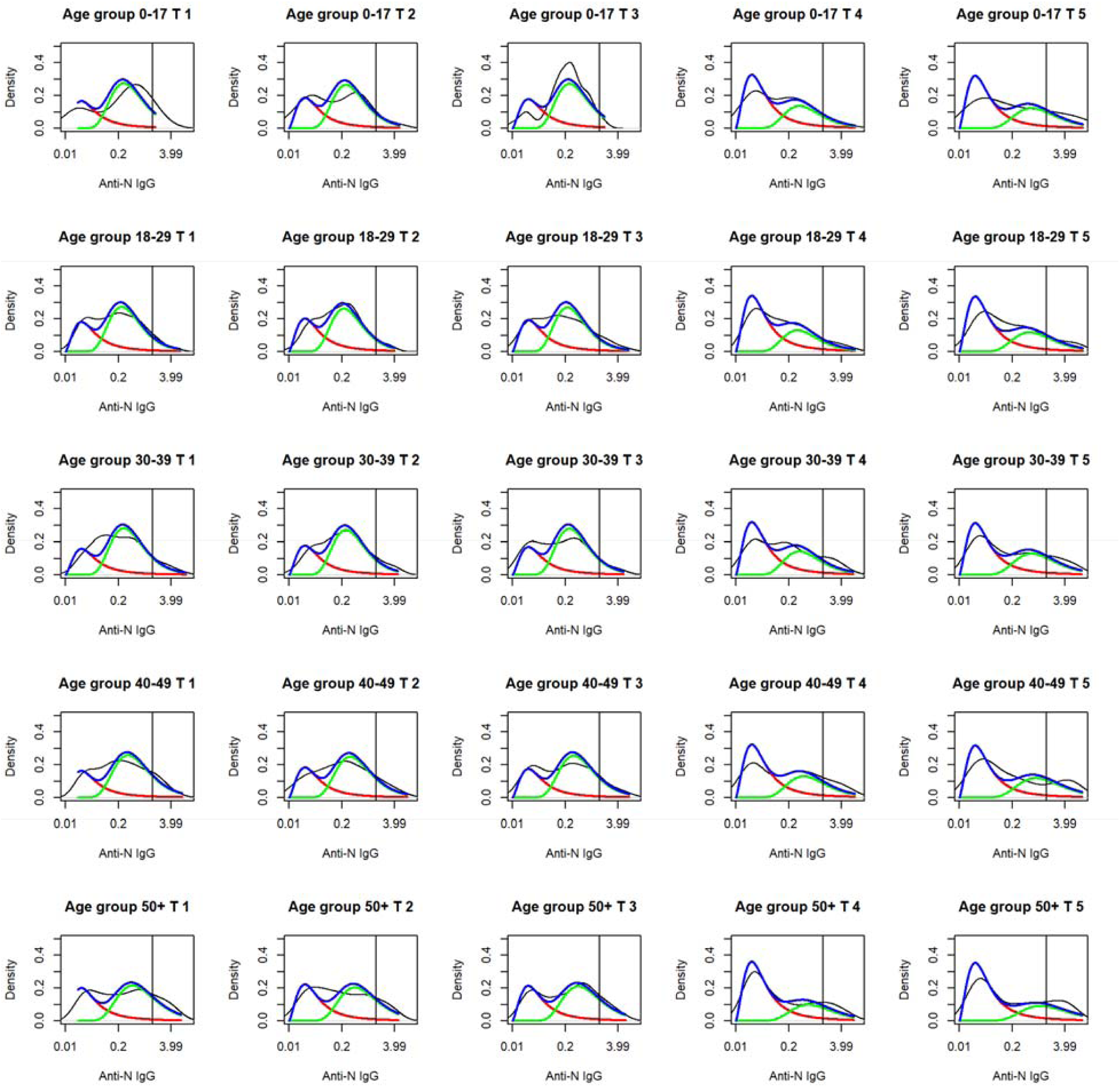
Null mixture model fit to anti-N IgG density by time and age quintile. Red lines represent the density of the seronegative component, green lines the density of the seropositive component, and the blue line represents the overall anti-N IgG density from the mixture model. The black lines represent the observed density. Vertical black lines represent the manufacturer’s cut-off.

**Fig. S3.**
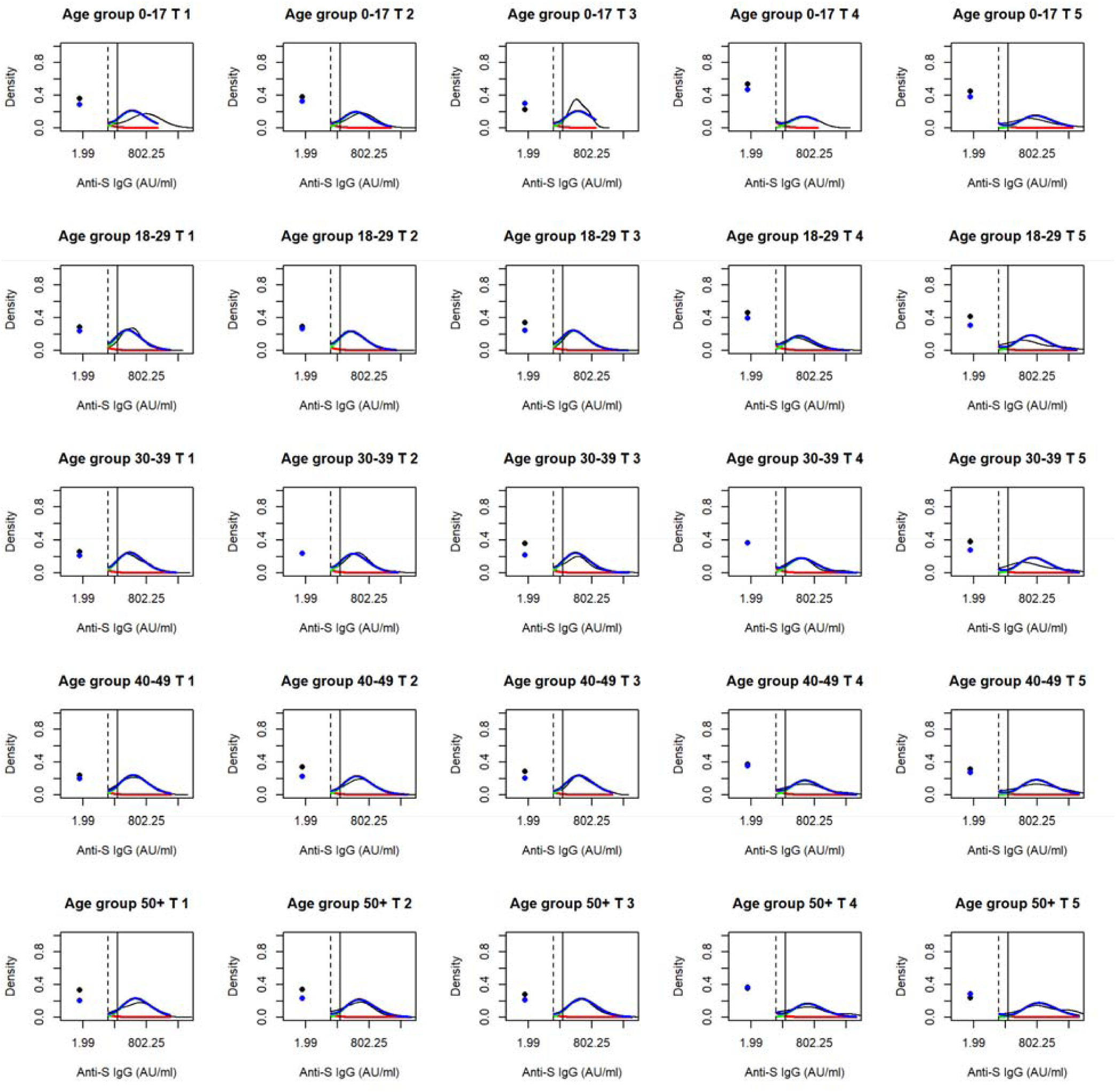
Null mixture model fit to anti-S IgG density by time and age quintile. Red lines represent the density of the seronegative component, green lines the density of the seropositive component, and the blue line represents the overall anti-N IgG density from the mixture model. Points represent the probability of being below the LLOQ, for the data (black) and the model (blue). The black lines represent the observed density. Vertical solid black lines represent the manufacturer’s cut-off, and vertical dashed black lines represent the LLOQ.

**Fig. S4.**
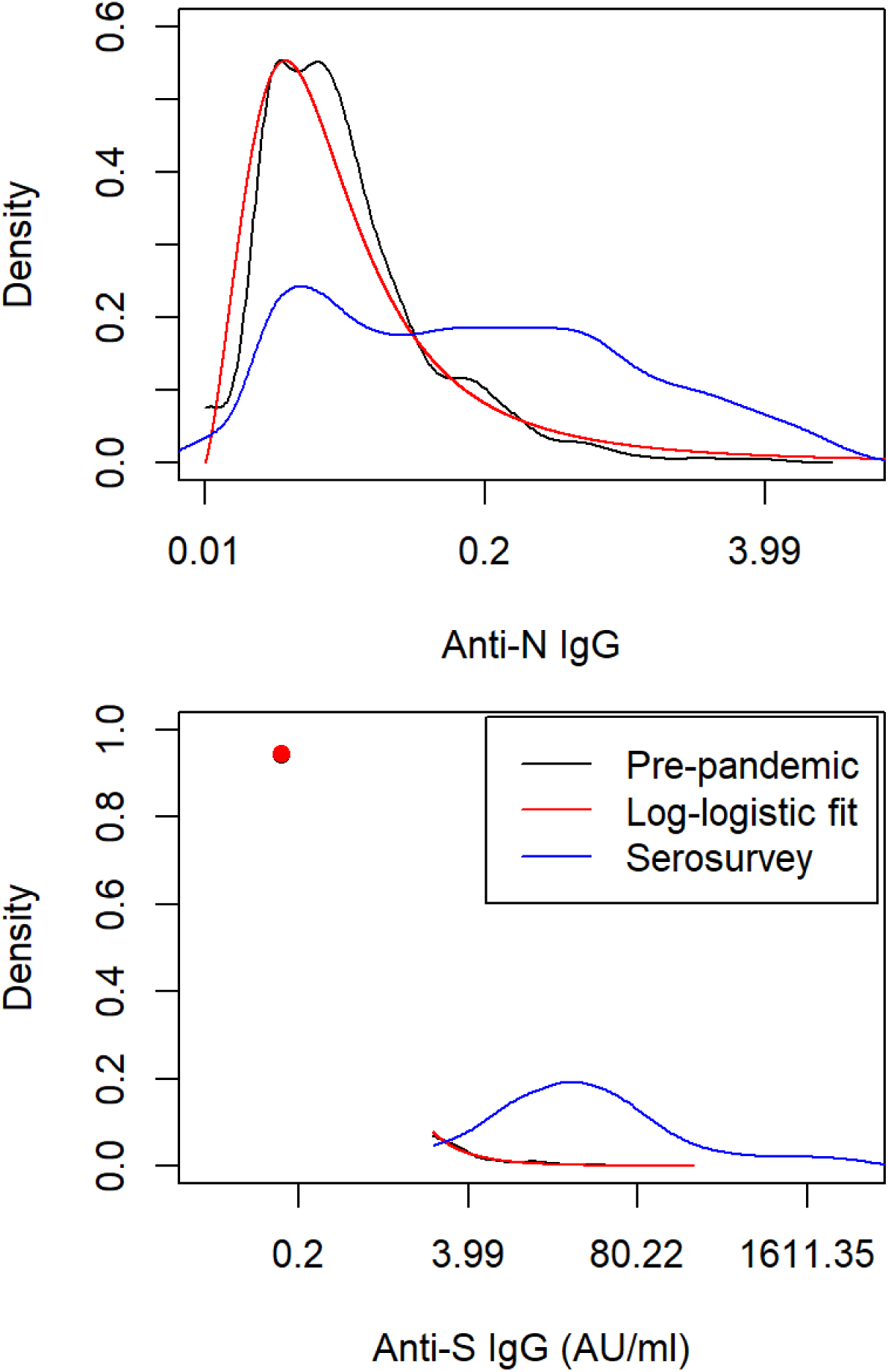
Mixture model fit to pre-pandemic seronegative samples, with the density of IgG in black, the model fit in red, and IgG density from the serosurvey in blue. For anti-S IgG, the points represent the probability of being below the LLOQ in the sample (black) and estimated from the model (red)

**Fig. S5.**
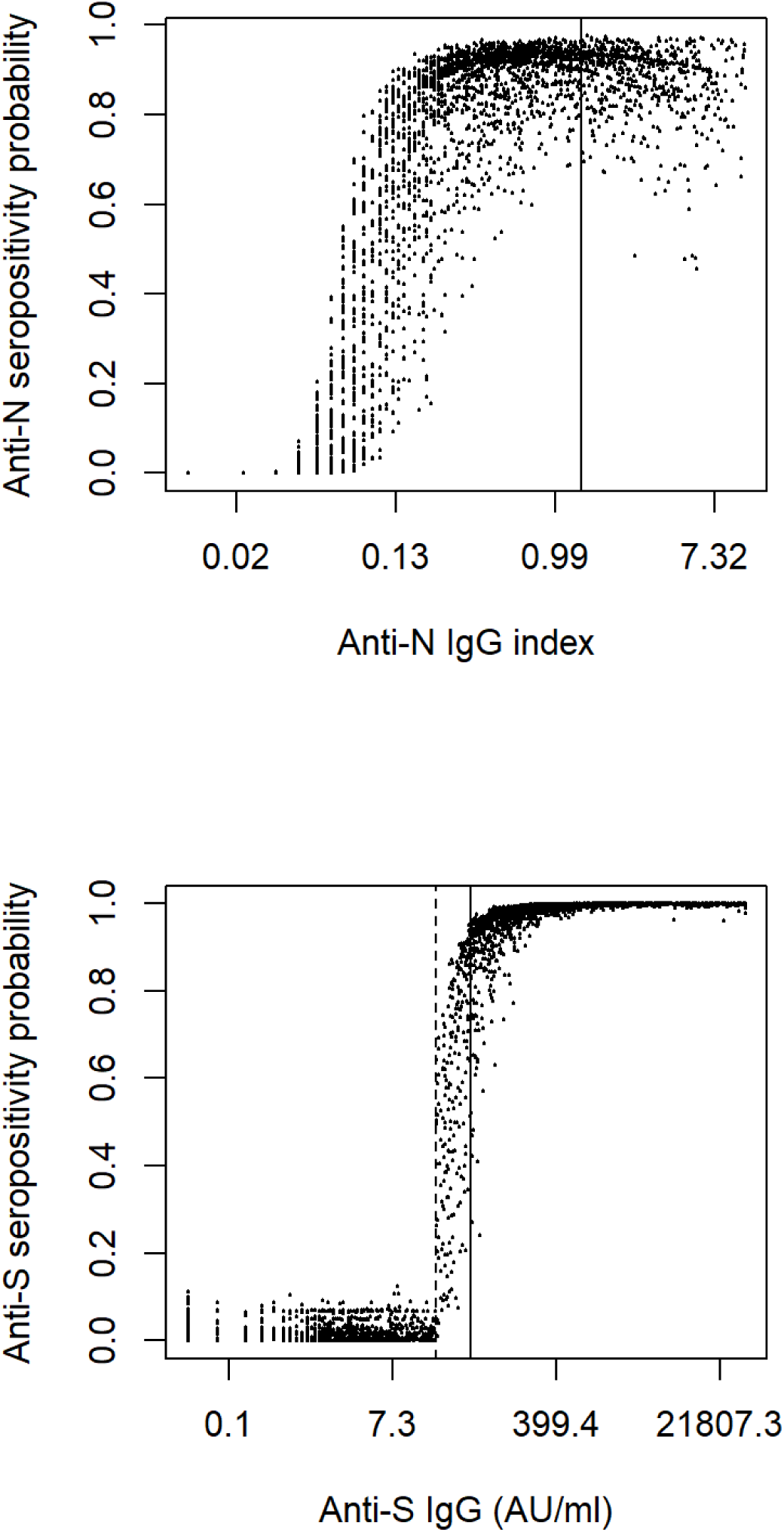
Model-estimated seropositivity probability for anti-N (top) and anti-S (bottom) IgG against measured IgG. Vertical solid lines represent the manufacturer’s cut-offs for each assay, and the vertical dotted line represents the LLOQ for the anti-S IgG assay

**Supplementary Table 1.** Anti-S IgG seroprevalence relative to anti-N IgG seroprevalence among individuals who did not self-report receipt of a COVID-19 vaccine, over time and by age group

|  | anti-S IgG seroprevalence / anti-N IgG seroprevalence (95% CrI) |
| --- | --- |
| Age (years) |  |
| 5-17 | 1.10 (1.04,1.18) |
| 18-29 | 1.22 (1.16,1.29) |
| 30-39 | 1.18 (1.13,1.24) |
| 40-49 | 1.18 (1.13,1.24) |
| ≥50 | 1.22 (1.17,1.27) |
| Time |  |
| 9 Jan – 14 Feb | 1.15 (1.10,1.20) |
| 15 Feb – 9 Mar | 1.15 (1.10,1.20) |
| 10 Mar – 24 Mar | 1.18 (1.13,1.24) |
| 25 Mar – 19 Apr | 1.28 (1.20,1.38) |
| 20 Apr – 13 May | 1.34 (1.26,1.44) |

\*CrI, Credible interval; N, Nucleocapsid; S, Spike

**Supplementary Table 2.** Difference in mean seroresponse to anti-N and anti-S SARS-CoV-2 IgG among SARS-CoV-2 seropositive individuals by age and sampling time

|  | Difference in log anti-N IgG index<br>among seropositive (95% CrI) | Difference in log anti-S IgG AU/ml<br>among seropositive (95% CrI) |
| --- | --- | --- |
| Age (years) |  |  |
| 5-17 | 0.13 (-0.08,0.36) | 0.45 (0.21,0.69) |
| 18-29 | REF | REF |
| 30-39 | 0.13 (-0.03,0.28) | 0.22 (0.07,0.37) |
| 40-49 | 0.39 (0.23,0.54) | 0.55 (0.39,0.71) |
| ≥50 | 0.68 (0.52,0.84) | 0.78 (0.63,0.92) |
| Time of sampling |  |  |
| 9 Jan – 14 Feb | 0.09 (-0.06,0.24) | -0.09 (-0.25,0.06) |
| 15 Feb – 9 Mar | REF | REF |
| 10 Mar – 24 Mar | 0.00 (-0.17,0.16) | -0.05 (-0.22,0.12) |
| 25 Mar – 19 Apr | 0.40 (0.20,0.59) | 0.30 (0.11,0.48) |
| 20 Apr – 13 May | 0.93 (0.72,1.14) | 1.14 (0.95,1.32) |

\*CrI, Credible interval; N, Nucleocapsid; S, Spike

